# Resilience and Social Support as Predictors of Life Satisfaction in Healthy Older Adults: A Moderation Analysis

**DOI:** 10.1101/2025.05.24.25328287

**Authors:** Lorena González, Aranzazu Duque, Mireia Molins, Marta Aliño, Francisco Molins, Patricia Mesa-Gresa

## Abstract

Previous literature has highlighted the importance of psychosocial factors such as the perception of an adequate social support network or the subjective experience of life satisfaction in promoting well-being in older adults and facilitating healthy aging processes. The objective of the present study was to analyze the possible relationship between the level of resilience and social support in healthy older adults, and how these variables may influence experiences of life satisfaction. Furthermore, potential moderation relationships between these variables were analyzed. The sample consisted of 42 healthy older adults (71.4% women), with a mean age of 66.57 years (SD = 5.82). Participants completed a battery of questionnaires that included a sociodemographic questionnaire, the Connor-Dadson Resilience Scale (CD-RISC-10), the Functional Social Support Questionnaire (Duke-UNC-11), and the Satisfaction with Life Scale (SWLS). The results of the multiple regression analyses indicate that resilience and social support positively and significantly predict life satisfaction. Furthermore, moderation analyses indicate that social support moderates the relationship between resilience and life satisfaction, jointly explaining 44.98% of the variance of this indicator of well-being. These results reveal that resilience in healthy older adults is related to their experiences of life satisfaction, and that the social support they perceive in their environment moderates this relationship. This underscores the need to expand the study of the correlations of well-being in old age, as well as to develop intervention programs aimed at promoting optimal aging that include strategies to improve resilience and foster functional social support networks.

## INTRODUCTION

Population aging represents a complex demographic phenomenon of great relevance in the 21st century and a global challenge, with significant social, health, economic, and population-related consequences (Wang, 2020; WHO, 2024). To address the challenges resulting from the rapid aging of the population, it is essential to investigate which factors may contribute to the promotion of healthy aging, enabling older adults to enjoy adequate cognitive functioning and greater well-being and quality of life (Krivanek et al., 2021). In this regard, scientific literature highlights the importance of studying how various psychosocial factors—such as social support, personality, or health status—can influence well-being and life satisfaction during this stage of life (Chiu et al., 2023; Chowdhary et al., 2022; Taylor & Carr, 2020).

Gaining a better understanding of the situation of older adults requires recognizing the significant and frequent transformations experienced during this stage of life, including changes in health status, retirement, the loss of significant others, or changes in financial situation. All these changes can lead to a deterioration of emotional well-being, contributing to sleep disturbances, dysregulation of the stress response, and disorders such as anxiety and depression, which in turn worsen quality of life and cognitive performance, and represents risk factor for the development of dementia (Chételat et al., 2018). Furthermore, during the later years of life, people often experience increased isolation and loneliness, which are associated with greater sedentarism, fewer health-promoting behaviors, and a higher prevalence of mood disorders such as anxiety or depression (Casamitjana & Cuevas, 2023).

The changes described above may lead to a decline in subjective well-being during this stage of life. Among the most studied indicators of well-being in the older population, life satisfaction stands out (Madhava Chandran et al., 2024; Tian & Chen, 2022), defined as the global and subjective cognitive evaluation of the extent to which an individual is satisfied with their life based on their goals and achievements (Diener et al., 1985). The concept of life satisfaction involves assessing and comparing actual life circumstances to expected ones (Kjell et al., 2016). When considering the evolution of life satisfaction across the lifespan, it is commonly believed that it gradually declines with age due to increased dependency, health problems, the loss of close relationships, or the proximity of death (Baumann et al., 2020). However, some studies have argued that life satisfaction and happiness increase with age (An et al., 2020). Factors influencing life satisfaction in older adults include health status, economic level, social support, type of pension, and intergenerational support (Tian & Chen, 2022). Previous studies analyzing the correlates of life satisfaction in late adulthood highlight the negative association between life satisfaction and the risk of chronic illness and mortality (Rosella et al., 2019; Lin et al., 2024), as well as the relationship between life satisfaction and healthy aging (Sone et al., 2022). All this evidence underscores the importance of exploring the factors that contribute to life satisfaction in older adults and how they interrelate.

One of the personality traits most frequently associated with healthy aging and subjective well-being in older adults is resilience (Trică et al., 2024), understood as the ability to quickly and effectively cope with and recover from difficulties, stress, and adversity through adaptation (Chen et al., 2024; Kais & Islam, 2016; Troy et al., 2022). Positive psychology emphasizes the importance of resilience in promoting mental health adaptations (Bartley et al., 2024). It is suggested that in adverse situations, highly resilient individuals are expected to adapt to and recover from setbacks and difficult experiences more easily (Harvanek et al., 2021; Zábó et al., 2023; Zeng et al., 2024). Studies focused on older adults have confirmed that those with higher levels of resilience appear to have greater capacity to thrive in the face of adversity or disruptive events (Chan et al., 2022; Gijzel et al., 2019). Research on the relationship between resilience and life satisfaction has shown that increased resilience predicts greater life satisfaction (Guo, 2017; Shabani et al., 2023). In fact, resilience is positively and significantly correlated with higher life satisfaction throughout the lifespan (Trică et al., 2024; Zheng et al., 2020), and especially among older adults (Liao et al., 2022). Resilient older adults are considered to have greater flexibility, higher confidence, longer life expectancy, more capacity to forgive others, a greater sense of purpose, more social participation, and a more positive outlook on life and the future (Hiebel et al., 2021). However, although resilience is a key individual factor in promoting life satisfaction, it seems that its role could be greatly influenced by the context in which people live. In this sense, some authors highlight the importance of a good social support network both for maintaining resilience (Górska et al., 2022) and for promoting well-being in older adults (Park & Sok, 2020; Zheng et al., 2020).

Focusing on the role that social support plays in promoting well-being in old age, experts have long recognized the importance of this contextual factor in the prediction of life satisfaction in older adults (Bai et al., 2018; Khodabakhsh, 2021; Li et al., 2018; Park & Sok, 2020; Zhang et al., 2017; Zheng et al., 2020). According to Cohen et al. (2020), social support includes the psychological and physical resources provided by social networks that help individuals cope with stress and negative moods. Social support has been shown to be associated with improvements in mental health (Acoba, 2024), and people who receive support from family, friends, or professionals tend to report higher levels of happiness and life satisfaction (Cao & Zhou, 2021) and higher resilience (Górska et al., 2022). Along these lines, Zhou et al. (2022) propose that social support intervenes in the relationship between resilience and life satisfaction. Thus, given the importance of this contextual factor, intervention programs have been developed that foster social support networks and consequently strengthen resilience and well-being in older individuals (e.g., Czaja et al., 2018; Liddle et al., 2024).

The findings to date underscore the relevance of personal factors, such as resilience, and social factors, such as perceived social support, for maintaining adequate mental health and improving experiences of life satisfaction and other well-being indicators. However, there is still limited literature that jointly examines the role of both resilience and social support in predicting life satisfaction among older adults. Moreover, the studies conducted to date show a variety of findings regarding the role of social support and resilience in predicting well- or ill-being. For instance, Zheng et al. (2020) found that older adults’ resilience partially mediates the relationship between perceived filial support and life satisfaction. While studies conducted in other populations have found that perceived social support mediates the relationship between resilience and quality of life in women with breast cancer (Zhou et al., 2022), and that perceived social support mediates the relationship between resilience and burnout in caregivers of older adults (Ong et al., 2018).

The state of the art, as well as the importance of life satisfaction in promoting healthy aging, highlights the need for further exploration of the role of resilience and perceived social support in predicting well-being. Thus, the general aim of the present study was to analyze the relationship between resilience, perceived social support, and life satisfaction in healthy older adults. To this end, the first specific objective focused on determining the potential predictive value of resilience and social support on life satisfaction. The second specific objective aimed to analyze the potential moderation effect of social support on the resilience-life satisfaction relationship among older adults. Results obtained through this work may help to clarify the relationship between the studied variables and may serve as a foundation for designing interventions aimed at strengthening psychosocial resources in older adults, thereby promoting healthier and more satisfying aging.

Based on the aforementioned specific objectives and current literature, the following hypotheses were proposed: (H1) Resilience and perceived social support will positively predict life satisfaction (Khodabakhsh, 2021; Shabani et al., 2023); and (H2) Social support will moderate the relationship between resilience and life satisfaction in older adults (Ong et al., 2018; Zhou et al., 2022).

## Method

### Participants

The study participants were 42 older adults, aged between 57 and 83 years (mean age = 66.57; *SD* = 5.82), with 71.4% of the sample being women. Participants were recruited using convenience sampling conducted within a university program for older adults. All participants met the following inclusion/exclusion criteria: being over 55 years of age, not presenting sensory or motor problems that would prevent them from completing the questionnaires and exceeding the Mini-Mental State Examination (MMSE) Questionnaire cutoff point (25 points). This study was conducted in accordance with the Declaration of Helsinki and was approved by the Ethics Committee of the university where the study was conducted (Reference: 2023-PSILOG-2558999).

### Instruments

**Sociodemographic Questionnaire.** It was developed *ad hoc* for this study. The information collected includes variables such as gender, age, educational level, marital status, number of children, and number of people living with.

**Mini-Mental State Examination Questionnaire** (MMSE; Folstein et al., 1975. Spanish adaptation by Lobo et al., 1995). This questionnaire is extensively used in clinical and research settings to measure cognitive impairment, including simple tasks in a number of areas: the test of time and place, the repeating lists of words, arithmetic such as serial subtractions of seven, language use and comprehension, and basic motor skills. It consists of 30 items, with a maximum score of 30 points; lower scores indicate greater cognitive impairment. The study validation reported a sensitivity of 0.85, a specificity of 0.90, and an intra-rater reliability of 0.93 (Lobo et al., 1999).

**Connor-Davidson Resilience Scale** (CD-RISC-10; Connor & Davidson, 2003. Spanish adaptation by Borche-Pérez et al., 2012). This scale assesses resilient behaviors in adults. It consists of 10 items answered using a five-point Likert scale ranging from 0 “*not at all*” to 4 “*almost always*”. An example item is “I am able to adapt when changes occur”. Responses can be grouped into a single dimension and indicate that higher scores suggest greater resilience. This scale has been used in other studies, showing a reliability of α = 0.87 (Soler Sánchez et al., 2016).

**Functional Social Support Questionnaire** (Duke-UNC-11; Broadhead et al., 1988. Spanish adaptation by Bellón et al., 1996). This instrument assesses perceived functional social support through 11 items grouped into two dimensions: confidential support (the ability to communicate with others) and affective support (demonstrations of love, affection, and empathy). An example of an item for the confidential support dimension is “I have the opportunity to talk to someone about my personal and family problems”, and an example of an item for the affective support dimension is “I receive love and affection”. Items are answered using a five-point Likert scale ranging from 1 “*Much less than you would like*” to 5 “*As much as you would like*”. Scores range from 11 to 55 points, with higher scores indicating greater perceived social support. In the Spanish validation, a score of 32 or lower indicates a low level of perceived social support, while a score above 32 indicates a normal level of perceived social support. Previous studies have demonstrated adequate internal consistency of the instrument (global evaluation: α = 0.89; confidential support dimension: α = 0.87; affective support dimension: α = 0.74) (Cuéllar-Flores & Dresch, 2012).

**Satisfaction with Life Scale** (SWLS; Diener et al., 1985. Spanish adaptation by Atienza et al., 2000). This scale is used to assess psychological well-being and quality of life by analyzing global cognitive judgments regarding the life of the person being evaluated. It is composed of 5 items answered using a seven-point Likert scale ranging from 1 “*Strongly disagree*” to 7 “*Strongly agree*”. An example item is “In most aspects my life is the way I want it to be”. Previous studies have shown good internal consistency of the instrument (α = 0.86) (Ramírez Pérez & Lee Maturana, 2012).

### Procedure

The study sessions took place from Monday to Thursday between 9:00 AM and 12:45 PM and lasted approximately 45 minutes to 1 hour. Participants were contacted and arranged to meet at a designated location at the faculty where the study was conducted. Once in the laboratory, they were explained the general procedure, and they read and signed the corresponding informed consent form. In addition, they received a brief assessment of their cognitive status using the Mini-Mental State Examination (MMSE) Questionnaire. Participants then completed questionnaires assessing the variables of interest.

### Analysis

Outliers were identified using the 2.5 standard deviation method. Normality was assessed via the Kolmogorov-Smirnov test with Lilliefors correction. Exploratory analyses were conducted to investigate the distribution of variables and their interrelationships. Subsequently, regression analyses were performed to determine whether resilience and social support could predict life satisfaction and to evaluate which of the two variables had greater predictive value. Finally, moderation analyses were conducted to further explore the dynamic between resilience and life satisfaction, incorporating social support as a moderator. The Johnson-Neyman procedure was applied to examine these interactions in greater depth. The significance level (α) was set at .05, and partial eta squared (η²p) was used to indicate effect size. All analyses were conducted using IBM SPSS Statistics 25.

## Results

### Descriptive Statistics

The general descriptive statistics for the sample are presented in Table 1. These variables were compared by gender to assess potential differences between men and women. As shown in Table 1, no significant differences were observed in any of the measured variables. Although life satisfaction scores indicated a trend toward significance, with men reporting slightly higher satisfaction than women, this did not reach statistical significance (*p* = .059). It is important to consider the disproportionate gender distribution in the sample (71.43% women and 28.57% men), which necessitates caution when interpreting these results, as this imbalance could increase the likelihood of a Type II error.

**Table 1-.** Descriptive statistics of the scores obtained in the CD-RSC-10 (resilience), Duke-UNC-11 (social support) and SWLS (satisfaction with life) questionnaires & gender comparison.

| | Global<br>(N = 42) | Men<br>(N = 12) | Women<br>(N = 30) | F | df<br>between | df<br>within | p-<br>value | $\eta^2_p$ |
| --- | --- | --- | --- | --- | --- | --- | --- | --- |
| Age | 66.57 ± 5.82 | 66.33 ± 5.17 | 66.67 ± 6.14 | 0.02 | 1 | 40 | .860 | .001 |
| Resilience | 73.24 ± 13.67 | 78.83 ± 9.39 | 71.00 ± 14.58 | 2.94 | 1 | 40 | .094 | .069 |
| Social support | 44.12 ± 7.69 | 43.92 ± 9.02 | 44.20 ± 7.27 | 0.11 | 1 | 40 | .916 | .000 |
| Life satisfaction | 27.31 ± 5.32 | 29.75 ± 3.64 | 26.33 ± 5.62 | 3.76 | 1 | 40 | .059<sup>†</sup> | .086 |

Mean ± standard deviation. Men and women were contrasted through ANOVA. A trend toward significance is indicated by <sup>†</sup> ( $p < .07$ ).

### Interrelation between Resilience, Social Support & Life Satisfaction

The analysis of Pearson correlations among the key variables indicated that resilience was positively correlated with life satisfaction, *r*(40) = .409, *p* = .007, suggesting that higher levels of resilience are associated with greater life satisfaction. Similarly, social support was also positively correlated with life satisfaction, *r*(40) = .421, *p* =.006, indicating that higher social support is linked to greater life satisfaction. However, there was no significant correlation between resilience and social support, *r*(40) = .108, *p* = .497. These findings suggest that while both resilience and social support individually relate to life satisfaction, they do not significantly correlate with each other within this sample.

### Regression analyses

To determine whether resilience and social support could predict life satisfaction, a multiple linear regression analysis was conducted. The predictors included in the model were total scores of resilience and social support, with life satisfaction as the dependent variable. The overall regression model was significant, *F*(2,39) = 8.805, *p* = .001, explaining 27.6% of the variance in life satisfaction (R^2^ = 0.27, with an adjusted R^2^ = 0.311). In line with the correlation analyses, social support emerged as a significant positive predictor of life satisfaction (*B* = 0.264, *SE* = 0.092, *t* = 2.851, *p* = .007), indicating that higher levels of social support are associated with greater life satisfaction. Similarly, resilience was also a significant predictor (*B* = 0.143, *SE* = 0.052, *t* = 2.754, *p*=.009), suggesting that higher resilience contributes to increased life satisfaction. These results affirm that both social support and resilience are important factors in predicting life satisfaction during aging. While both predictors were statistically significant, the slightly higher beta coefficient for social support suggests it may have a stronger influence on life satisfaction than resilience.

### Moderation analyses

Building on the insights from the previous regression analyses, a moderation analysis was performed to further explore the dynamic between resilience and life satisfaction, incorporating social support as a moderator. This analysis aimed to dissect the nuances not captured by the initial regression model, which already indicated significant roles for both resilience and social support in predicting life satisfaction.

The comprehensive model confirmed the significant predictive power of these variables, explaining approximately 44.98% of the variance in life satisfaction (*F*(3,38) = 10.354, *p* < .001). Resilience continued to show a positive effect on life satisfaction (*B* = 0.765, *SE* = 0.206, *t* = 3.708, *p* = .001), a finding consistent with earlier results. Social support also maintained its significant positive impact (*B* = 1.314, *SE* = 0.350, *t* = 3.759, *p* < .001). However, the interaction between resilience and social support introduced a new layer of complexity (*B* = −0.014, *SE* = 0.005, *t* = −3.095, *p* = .004), indicating a moderating effect. This interaction significantly altered the relationship between resilience and life satisfaction, contributing an additional 13.87% to the explained variance (R^2^change = 0.139, *F*(1,38) = 9.580, *p* = .004).

The Johnson-Neyman technique was utilized to pinpoint where social support levels shift the influence of resilience on life satisfaction. The results indicated that the effectiveness of resilience in enhancing life satisfaction becomes notably significant only when social support scores fall below 47.3517 (see Figure 1). This threshold highlights a distinct divergence from the simpler effects modelled in the multiple regression, where the influence of individual predictors was not conditioned on the level of social support. In regions where social support exceeds this critical value, the relationship between resilience and life satisfaction does not manifest significantly. Conversely, with lower social support, this relationship not only emerges but also strengthens, underscoring the pivotal role of social environments in leveraging personal strengths like resilience.

**Figure 1.**
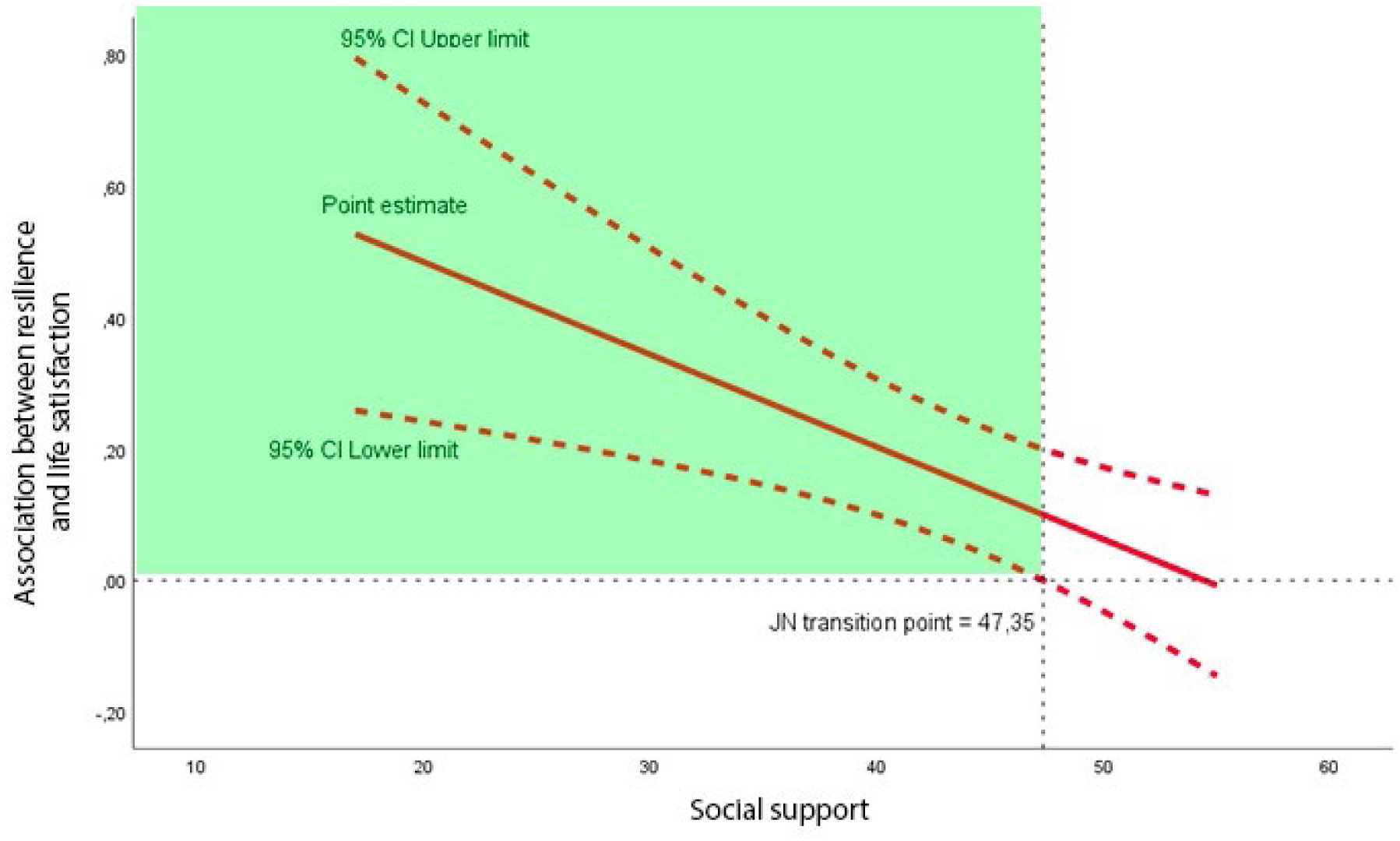
Johnson-Neyman graph. This graph illustrates the conditional association between resilience and life satisfaction as a linear function of social support. The plot includes the Johnson–Neyman (JN) transition point, which marks the value of social support at which the effect of resilience on life satisfaction becomes statistically significant. This point corresponds to where the confidence interval for the conditional effect intersects zero on the Y-axis. The green shaded area represents the region of significance—that is, the range of social support values for which the association between resilience and life satisfaction is significant.

In summary, the moderation analysis reveals that the impact of resilience on life satisfaction in a sample of healthy older people is contingent upon the level of social support, an insight that extends beyond the direct effects observed in the initial regression models. This nuanced understanding suggests that enhancing social support could be a key strategy in maximizing the positive effects of resilience on life satisfaction.

## Discussion

The main objective of this study was to analyze the relationship between resilience, perceived social support, and life satisfaction in healthy older adults. The overall results indicate that both resilience and the perceived social support of older individuals positively and significantly predict their life satisfaction. Additionally, the results show that perceived social support moderates the relationship between psychological resilience and life satisfaction, with resilience becoming a stronger predictor when perceived social support is lower. In fact, the results suggest that among older adults with high social support, the predictive power of resilience weakens and predicts life satisfaction to a lesser extent; however, as perceived social support decreases, the relationship between resilience and life satisfaction becomes closer. These findings highlight the crucial role of an adequate social support network in fostering subjective well-being among older adults and emphasize how important resilience becomes when social support is lacking.

Based on the descriptive statistics of the study, participants perceive themselves as highly resilient, with good social support and a high level of life satisfaction. It is also noteworthy that these results are consistent across both men and women participants, with no significant gender differences found in the variables assessed. These results are in line with previous studies that have reported that older adults generally indicate moderate to high levels of resilience (Treichler et al., 2020; Zeng et al., 2024; Zheng et al., 2020), perceived medium to high social support (Park & Sok, 2020; Zheng et al., 2020), and high life satisfaction (Tian & Chen, 2022; Zheng et al., 2020). Moreover, several systematic reviews have emphasized that no gender differences are typically found in life satisfaction among older people (Cheng et al., 2021), and that gender differences in resilience perception are either minimal or inconsistent (Górska et al., 2022). Regarding social support, previous research has found no gender differences in the perceived social support among older adults, although gender differences were found in perceived instrumental support (Abu-Kaf et al., 2022). While these results depict a relatively positive situation for older adults, it is important to point out that most studies, including the present one, have involved healthy, community-dwelling older adults, so differences could be expected if the study were conducted with individuals experiencing aging with serious illnesses.

Regarding the proposed hypotheses in the present study, the first one suggested that resilience and perceived social support will positively predict life satisfaction experienced by older adults. The results in the present study confirm these relationships and indicate that when older adults feel resilient and capable of coping with adversity, this leads to greater life satisfaction. Studies involving different age groups suggest that those with higher resilience also report greater life satisfaction, possibly due to the personal resources they possess for coping with challenges (Yang et al., 2020; Guo, 2017). Specifically, in older adults, evidence suggests that resilience contributes holistically to healthy aging and is positively related to life satisfaction and other well-being indicators such as quality of life, optimism or positive emotions (Górska et al., 2022; Zeng et al., 2024; Zheng et al., 2020). On the other hand, the results of the present study show that when older individuals perceive themselves as having a strong social support network —comprised of people they trust and who offer them affection and care— they experience greater life satisfaction. These findings align with previous research showing that social support is associated with higher life satisfaction, both in older adults (Park & Sok, 2020; Zheng et al., 2020) and in other populations (Cao & Zhou, 2019). Even more, comparing the effect of both independent variables on life satisfaction, the results of this study indicate that perceived social support has a greater impact on life satisfaction than resilience. Although previous literature, both in older populations and other groups, does not show a consistent trend in these relationships (Ong et al., 2018; Zheng et al., 2022), these results underscore the importance of providing resources that help older adults build resilience and strengthen their social support networks, as this contributes to greater well-being.

Regarding the second hypothesis, it was proposed that perceived social support in older adults could act as a moderator in the relationship between resilience and life satisfaction. The present results support this hypothesis, showing that the direct effect of resilience and the moderating role of social support together explain 44.98% of the variance in life satisfaction among older adults. Furthermore, the analysis revealed that the impact of resilience on life satisfaction varies depending on the level of perceived social support. Specifically, as perceived social support increases, the predictive value of resilience decreases. Along the same line, previous studies already found that social support exerted an important role in the relationship between resilience and variables such as quality of life (Zhou et al., 2022), caregiver burnout (Ong et al., 2018), or sleep quality (Cui et al., 2024). These studies support the idea that social support can be conceived as a protective factor for older adults (Cihlar et al., 2022). Research conducted with other populations has also demonstrated associations between resilience, perceived social support, and subjective well-being (Yang et al., 2024), as well as with distress-related outcomes such as anxiety or depression in clinical populations (Sippel et al., 2015). It is important to note that some recent studies have approached these variables differently, finding that resilience partially mediated the relationship between social support and life satisfaction in older adults (Zheng et al., 2020). These findings provide additional evidence of the crucial role that psychosocial factors play in promoting well-being and mitigating distress. However, due to the limited literature focused on older populations and the inconsistencies in existing findings, further research is needed to clarify the roles of resilience and social support in enhancing life experiences among the elderly.

As the world’s population ages, the psychological and physical health of elderly people has become a common concern of the global community. The results of this study have important implications for promoting well-being in older adults by identifying two key aspects that can foster life satisfaction: on one hand, the capacity for resilience to overcome adversity and grow through critical situations, and on the other hand the social support provided by family, friends, or professionals who care for them and show them affection. The findings suggest that intervention programs aimed at promoting the well-being of older adults should focus on strengthening or expanding their social groups, facilitating the creation of new groups that can serve as sources of connection and support, as well as providing resources for assertiveness and problem-solving during this life stage. Recent studies on intervention programs for older adults—mainly implemented through technological platforms—have demonstrated their effectiveness in expanding social support networks and reducing isolation or loneliness in later life, while also improving perceived social support and overall well-being (Czaja et al., 2018). Other interventions have specifically focused on strengthening resilience in older adults and have been shown to also improve quality of life (Terkes et al., 2023). Regardless of whether the central focus of interventions is on promoting one outcome or another, it is important that interventions are provided as individualized and multicomponent as possible (Chowdhary et al., 2022).

Despite the contributions of this study, it is important to acknowledge some limitations. First, the characteristics of the sample must be considered, as it consisted of 42 healthy older adults, 71.4% of whom were women. However, the gender difference observed in this study may reflect the composition of the specific sample, which consisted of older adults enrolled in a university course in the field of Psychology—a field predominantly chosen by women in the country of study. Additionally, previous studies have shown that older women participate in these types of studies much more frequently than men (Czaja et al., 2018; Rae et al., 2025). Moreover, the participants were recruited from a university program for older adults attending health-related courses, which means that the sample likely had better health and life conditions than many other older adults. A similar situation has been observed in previous studies that also recruit healthy, community-dwelling older adults, where most participants tend to be in a positive situation in terms of health or social conditions (Rae et al., 2025). Future research would benefit from expanding the sample to include a larger number of older adults, representative of both genders, and incorporating individuals from different social backgrounds. Additionally, future studies could include objective measures to strengthen the findings. While the instruments used are validated and have high experimental utility (Ruiz-Comellas et al., 2021; Zheng et al., 2020; Zhou et al., 2022), the use of objective variables, such as electrophysiological and/or hormonal assessments, could provide more robust evidence for the conclusions.

In general, the results of this study highlight two major promoters of life satisfaction in older adults: resilience and social support. This shows that if we aim to help older people feel more satisfied with their lives, it is essential to foster and strengthen resilience resources while simultaneously improving their social support networks. Moreover, the present findings help to clarify the relationship between these variables, suggesting that social support moderates the relationship between resilience and life satisfaction in older adults. This approach not only aims to identify the elements that enhance well-being in older adults but also provides a foundation for developing individualized and multicomponent interventions that promote holistic health and improve quality of life.

## Data Availability

All data produced in the present study are available upon reasonable request to the authors

## Acknowledgments

Research reported in this publication was supported by MCIN/AEI/10.13039/501100011033/ and by “ERDF A way of making Europe” under grant number PID2022-138021OA-I00.

## Notes

### Competing Interest Statement

The authors have declared no competing interest.

### Funding Statement

This study was funded by MCIN/AEI/10.13039/501100011033/ and by 'ERDF A way of making Europe' under grant number PID2022-138021OA-I00.

### Author Declarations

Ethics committee of Universitat de Valencia (Spain) gave ethical approval for this work

